# Tapestry: A Single-Round Smart Pooling Technique for COVID-19 Testing

**DOI:** 10.1101/2020.04.23.20077727

**Authors:** Sabyasachi Ghosh, Ajit Rajwade, Srikar Krishna, Nikhil Gopalkrishnan, Thomas E. Schaus, Anirudh Chakravarthy, Sriram Varahan, Vidhya Appu, Raunak Ramakrishnan, Shashank Ch, Mohit Jindal, Vadhir Bhupathi, Aditya Gupta, Abhinav Jain, Rishi Agarwal, Shreya Pathak, Mohammed Ali Rehan, Sarthak Consul, Yash Gupta, Nimay Gupta, Pratyush Agarwal, Ritika Goyal, Vinay Sagar, Uma Ramakrishnan, Sandeep Krishna, Peng Yin, Dasaradhi Palakodeti, Manoj Gopalkrishnan

**Affiliations:** IIT Bombay; NCBS Bangalore; InStem Bangalore; Wyss Institute for Biologically Inspired Engineering at Harvard University; Department of Systems Biology, Harvard Medical School; Shop101; SASTRA University

## Abstract

The COVID-19 pandemic has strained testing capabilities worldwide. There is an urgent need to find economical and scalable ways to test more people. We present **Tapestry**, a novel quantitative nonadaptive pooling scheme to test many samples using only a few tests. The underlying molecular diagnostic test is any real-time RT-PCR diagnostic panel approved for the detection of the SARS-CoV-2 virus. In cases where most samples are negative for the virus, Tapestry accurately identifies the status of each individual sample with a single round of testing in fewer tests than simple two-round pooling. We also present a companion Android application **BYOM Smart Testing** which guides users through the pipetting steps required to perform the combinatorial pooling. The results of the pooled tests can be fed into the application to recover the status and estimated viral load for each individual sample.

NOTE: This protocol has been validated with in vitro experiments that used synthetic RNA and DNA fragments and additionally, its expected behavior has been confirmed using computer simulations. Validation with clinical samples is ongoing. We are looking for clinical collaborators with access to patient samples. Please contact the corresponding author if you wish to validate this protocol on clinical samples.

## Introduction

Currently, the primary method for COVID-19 testing uses viral RNA extraction followed by RT-qPCR amplification of a conserved region of the genome of the virus. The throughput and capacity for such testing is severely limited. Since COVID-19 can be transmitted from asymptomatic carriers, these testing bottlenecks have left states with the dilemma of either risking a free spread of the virus, or imposing severe lockdown and physical distancing measures with heavy economic costs.

One strategy many countries have adopted to increase testing capacity is to combine samples into pools that are tested together [1,2,3,4,5]. If such a pool is tested negative, all individual samples within the pool are declared free of SARS-CoV-2, whereas if it is tested positive then all individuals in that pool must be retested individually in a second round. While these strategies can augment testing capacities, they do not substantially improve throughput, since two rounds of testing are still needed to identify positive samples.

A challenge that such simple pooling strategies have to confront is whether to first pool samples and then perform RNA extraction on the pools, or whether to individually extract RNA from each sample and subsequently pool the RNA. If RNA extraction is done individually for each sample, then the requirement to run a positive PCR control on the RP gene (confirming RNA extraction worked) for every sample would nullify any gains from pooling. If RNA extraction is performed after pooling samples, then the second round of testing would require a new round of RNA extraction for all individual samples from positive pools, slowing down the process. Nonadaptive testing allows a way past this dilemma, by pooling samples, extracting RNA from pooled samples, and returning confirmed sample-level results in a single round of testing.

Here we introduce **Tapestry Pooling**, a novel pooling strategy that increases both capacity and throughput beyond simple pooling. Tapestry pooling gives confirmed results in a single round of testing by testing each sample thrice as part of three different pools. The pooling design is chosen in a combinatorial manner to make decoding possible. The technique is quantitative, based on ideas from compressed sensing. The decoding algorithm takes as input cycle threshold values from qPCR tests on the pools, and returns a result for each sample, along with an estimate of viral load if positive. The number of tests required with Tapestry Pooling compares favorably with the two-round poolings that are currently deployed in the field with comparable levels of sensitivity and specificity (See Table 1).

**Table 1.** Comparison of one-round Tapestry pooling to two-round Dorfman pooling. Tapestry not only increases throughput by providing results in one round but can also reduce the required number of tests.

| #Samples | Prevalence | 2-round Dorfman | 1 round Tapestry |
| --- | --- | --- | --- |
| 105 | 7.6% | 65 | 45 |
| 40 | 7.5% | 22 | 16 |
| 70 | 5.7% | 34 | 21 |
| 195 | 4.1% | 89 | 45 |
| 399 | 2.5% | 127 | 63 |
| 961 | 1.0% | 197 | 93 |
| 1140 | 0.2% | 102 | 20 |

The solved viral load for each sample has to satisfy a quantitative consistency check. The sum of viral loads of samples in a pool must tally with the measured cycle threshold of the pool, with some allowance for noise. This quantitative reconstruction allows more information to be extracted from positive tests than from binary group testing approaches which take into account only whether a test is positive or negative. Specifically, in the binary group testing situation a positive test for a pool with one known positive sample does not convey any information about other samples in that pool. This is not the case with Tapestry pooling where the quantitative values of the test and the sample can be compared to reveal if there might be other samples in the pool that are positive. As a result, Tapestry pooling continues to be viable not just with low (<4%) prevalence rates, but even with moderate prevalence rates (5% - 10%), in which regime simple pooling does very poorly.

Our algorithm can estimate the number of positive samples in the population from the number of tests that came out positive. Based on this sparsity estimator, the algorithm has a graceful failure mode in case the prevalence rate is far above the assumed prevalence rate for which the test was designed. In such a case, our algorithm will still maintain very high sensitivity, returning almost no false negatives. It will return a list of sure positives, and a list of suspected positives with an advice on how many of the suspected positives are in fact positive. This list-decoding approach makes our algorithm viable to be deployed with prevalence rates even as high as 10%.

We have designed an Android app **BYOM Smart Testing** to facilitate implementation of Tapestry Pooling in clinical laboratory settings. The app encapsulates all the complexity of our scheme, and presents a simple and easy to execute protocol to the testing lab, vastly increasing the deployability of this scheme. The app provides a sequence of instructions in the form of visual cues to aid and direct the pipetting of samples into appropriate pools. Once pooled tests are run, the cycle threshold values can be entered into the app which solves for and displays the list of positive and negative samples along with an estimate of viral loads.

In this work, we focus on PCR tests since it is currently the most widely used testing methodology for COVID-19. In principle Tapestry Pooling may also be applicable to other testing assays like serological antibody tests, both for COVID-19 testing and other purposes.

We first refined and validated Tapestry Pooling through computer simulations. We then validated Tapestry Pooling analytically in a lab setting with RNA and DNA fragments in blinded experiments. In cases where our simulations predicted that we would, with high likelihood, recover individual sample status, our experiments indeed successfully did so. We are currently testing Tapestry with clinical samples.

## Methods

### Theoretical background

Let the samples be numbered 1, 2, …, *n* and indexed by *i*. Let the viral load of the i^th^ sample be given by *x_i_*. Suppose the tests are numbered 1, 2, …, *m* and indexed by *j*. Let 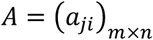 be a matrix with entries either 0 or 1. Then entry *a_ji_* = 0 means that sample *i* is not present in test *j*, and *a_ij_* = 1 means that sample *i* is present in test *j* (see Fig 1A). Let *y_j_* be the quantitative measure of viral load in the j^th^ test where *j* = 1, 2, …, *m*. Then 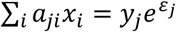 where *ε_j_* is a mean-zero Gaussian random variable denoting noise in measurement of cycle threshold for the j^th^ sample. Since most samples tested will not have the virus, we want to reconstruct a sparse (most entries 0) nonnegative vector x that satisfies this noisy linear equation. This problem has been studied in signal processing literature under the name of compressed sensing [7,8,9], though the combination of A being a sparse matrix with 0-1 entries, x being nonnegative, and the multiplicative noise model is unique to this particular situation and has prompted algorithmic innovations on our part.

**Fig 1.**
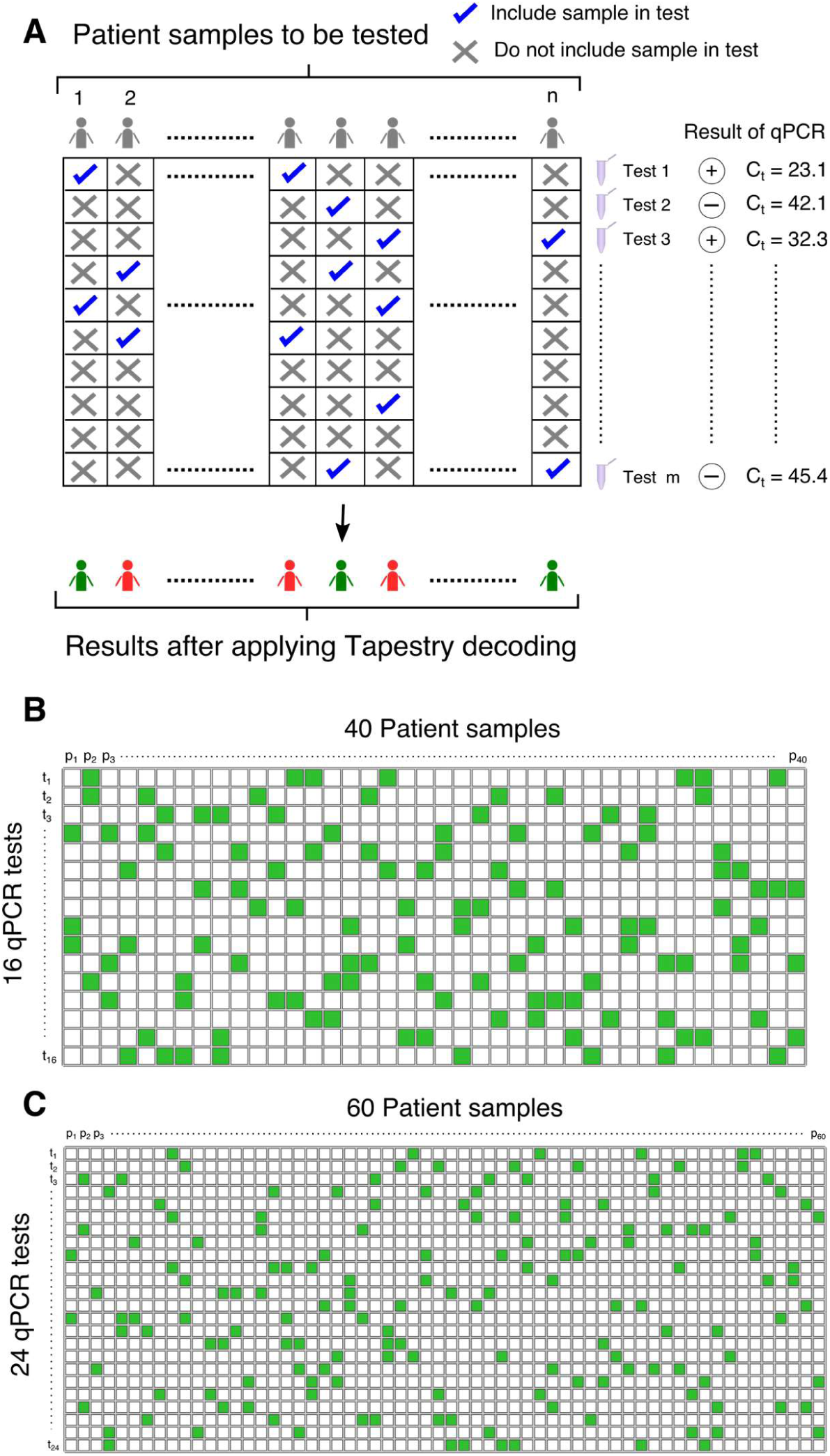
**A**. An overview of the tapestry pooling scheme. A total of n patient samples are pooled into m tests in a combinatorial manner, as indicated by the matrix. Each patient sample is included in multiple test and each test contains multiple patient samples. The result of each pooled test, in the form of a C_t_ value, is used to recover the status of individual samples. **B**. A 16 × 40 matrix which tests 40 patient samples in 16 tests that was analytically validated in vitro using mock samples made up of RNA fragments in a background of mouse RNA. **C**. A 24 × 60 matrix which tests 60 patient samples in 24 tests that was analytically validated in vitro using a mock sample made up of a circular DNA plasmid containing the complete nucleocapsid gene from the SARS-CoV-2 virus.

We have explored various ways to design the matrix A including sparse expanders, explicit optimization of a loss function in a gradient descent and simulated annealing fashion, and finally using Steiner triples. We find Steiner triples particularly powerful and convenient for this application. In matrices constructed from Steiner triples, each sample goes into 3 pools. This makes A a very sparse matrix which has advantages for pipetting and keeps pool sizes manageable. Further any two columns of A have dot product no more than 1 --- in other words each pair of samples occurs together in no more than one test. This property ensures good reconstruction guarantees for x. Further details of our decoding algorithms will be described in an upcoming theory paper.

### Experimental methods

#### RNA and DNA amplicons

A single stranded RNA fragment of length 1 kb (stock concentration 10 ng/µL) was used as a proxy for viral RNA for preliminary testing. RNA fragments were diluted to clinically relevant concentrations of 0.05 pg/µL (~10^5^ copies/µL), 5 fg/µL (~10^4^ copies/µL) and 0.5 fg/µL (~10^3^ copies/ µL) by serial dilution from the stock. We also tested our scheme with a circular DNA plasmid containing the complete nucleocapsid gene from the SARS-CoV-2 virus (Integrated DNA Technologies, 2019-nCoV_N_Positive Control, Catalog #10006625). DNA plasmid was diluted to clinically relevant concentrations of 10^4^ copies/µL and 10^3^ copies/µL.

#### Pooling

Two pooling matrices, a 16 × 40 (tests × samples) and a 24 × 60 (tests × samples), were tested in blinded experiments where the identity and viral load of the individual positive samples were not a priori revealed to the decoding team. Only the C_t_ values corresponding to the various pooled qPCRs were communicated.

The 16 × 40 matrix (see Fig 1B) directs that each sample be distributed, and hence tested, two or three times while the number of samples per pool vary from six to nine, with a median pool size of seven. The 16 × 40 matrix was tested for five different situations, corresponding to starting with zero positive samples, one positive sample (10^3^ copies), two positive samples (10^3^ copies each), three positive samples (two with 10^3^ and one with 10^4^ copies) or four positive samples (two with 10^3^, one with 10^4^ and one with 10^5^ copies). Sample pools containing one or more positive samples were emulated by spiking with appropriate amounts of RNA fragments to simulate the viral load after pooling. Mouse RNA (~ 160 pg per reaction) was added as background to simulate the expected clinical sample matrix. A total of 24 such sample pools were generated for downstream amplification. The many sample pools containing no positive samples were emulated by assuming that their C_t_ values were normally distributed with a mean of 33.85 and a standard deviation of 0.371, a distribution indicated by running qPCR experiments with no amplifiable templates, only background mouse RNA.

The 24 × 60 matrix (see Fig 1C) directs that each sample be distributed, and hence tested, two or three times while the number of samples per pool is either six or seven with a median pool size of seven. We tested a situation corresponding to two positive samples (10^4^ copies and 10^3^ copies of 2019-nCoV_N_Positive Control) and 58 negative samples (58 aliquots of nuclease free water in 58 separate tubes). The 60 mock samples were variously distributed to generate 24 sample pools for downstream amplification.

#### qPCR Amplification from RNA templates

cDNA was synthesized from RNA templates using First Strand Invitrogen SuperScript II system (Invitrogen, catalog #18064-014). A mix of RNA and a gene-specific reverse transcription primer were denatured at 95°C for 5 minutes and annealed at 50°C for 5 minutes prior to the addition of the other reaction components. After the other components were added, the reverse transcription reaction was carried out for 1 hour at 50°C and was followed by a heat inactivation step for 15 minutes at 85°C. For cDNA templates, qPCR was performed using the ThermoFisher SYBR GREEN MasterMix system (Thermo Scientific, catalog #K0223). A 35 µl qPCR reaction was set up using cDNA template (3.5 µL) and forward and reverse primers (2.5 µM each). Each 35µl reaction was distributed equally as 3 technical replicates of 10 µl in an optical 384 well plate as technical replicates. The thermocycling conditions were as follows: 95°C denaturation step for 3 minutes followed by 40 cycles of 95°C for 10 seconds, 55°C for 15 seconds and 72°C for 30 seconds. The specificity of the initial test reactions were verified by primer melt curve analysis and analysis of the amplicons using agarose gel electrophoresis.

#### qPCR Amplification from DNA templates

DNA templates were amplified with TaqPath 1-Step RT-qPCR Master Mix, CG (ThermoFisher; catalog #A15299) using the U.S. CDC designed N1 primer and Taqman probe set (IDT; 2019-nCoV RUO Kit, catalog #10006713). The RT step was skipped during thermocycling, as we started with a DNA template. The thermocycling conditions were as follows: 95°C denaturation step for 2 minutes followed by 45 cycles of 95°C for 3 seconds, 55°C for 30 seconds and 72°C for 30 seconds.

## Results

16 × 40 pooling matrix: Table 2 shows the emulated cycle threshold (C_t_) values for the 16 RT-qPCR tests corresponding to five different trials (0, 1, 2, 3 or 4 positive samples out of a total of 40 samples). Fig 2A shows the corresponding amplification curves. Table 3 shows the ground truth RNA amounts for each of the 40 samples and compares them to the values estimated using the Tapestry decoding algorithm. We find that for 0, 1 and 2 positives (for which simulations suggested the matrix would perform well with high likelihood) the algorithm correctly identified the status of all samples with zero false negatives and zero false positives. For 3 or 4 positives, as expected, our algorithm returned zero false negatives, but some false positives. However, all the false positives are correctly identified as having a low likelihood of being a true positive, and are placed in the set of unsure positives. All sure positive and sure negative sets returned by the algorithm were correct.

**Table 2.** Cycle thresholds (C_t_) values for five different pooling trials of the 16 × 40 pooling matrix from Fig 1B. Each column lists C_t_ values corresponding to a single pooling trial. See experimental methods section for details on experimental setup. Note that RT-qPCR was only performed for pools in which at least one sample contained amplifiable template. The corresponding amplification curves may be found in Fig. 2A. The many sample pools containing no positive samples were emulated by assuming that their C_t_ values were normally distributed with a mean of 33.85 and a standard deviation of 0.371, a distribution indicated by running qPCR experiments with no amplifiable templates. The algorithm was blinded to the status of the samples and only received 16 C_t_ values for each trial, and did not have any knowledge of which C_t_ values were from an actual an RT-qPCR run.

| 0 positive samples | 1 positive samples | 2 positive samples | 3 positive samples | 4 positive samples |
| --- | --- | --- | --- | --- |
| 33.73 | 31.60 | 33.49 | 34.05 | 31.31 |
| 33.87 | 33.72 | 33.91 | 33.55 | 30.64 |
| 34.10 | 33.63 | 30.93 | 26.17 | 33.57 |
| 33.52 | 33.71 | 33.85 | 34.06 | 33.85 |
| 33.95 | 30.47 | 34.00 | 31.02 | 26.23 |
| 34.19 | 34.04 | 33.50 | 30.07 | 27.43 |
| 33.95 | 33.65 | 34.09 | 34.13 | 34.10 |
| 34.11 | 34.04 | 31.04 | 27.54 | 26.38 |
| 33.56 | 33.79 | 30.54 | 34.14 | 33.75 |
| 33.51 | 33.75 | 34.18 | 31.16 | 33.77 |
| 34.15 | 34.20 | 33.70 | 34.10 | 22.68 |
| 33.69 | 33.94 | 34.21 | 26.33 | 33.86 |
| 33.76 | 33.83 | 33.96 | 34.01 | 33.82 |
| 33.53 | 30.73 | 33.50 | 34.10 | 22.57 |
| 33.95 | 33.69 | 30.67 | 33.83 | 33.53 |
| 33.97 | 33.97 | 30.17 | 30.63 | 22.71 |

**Fig 2.**
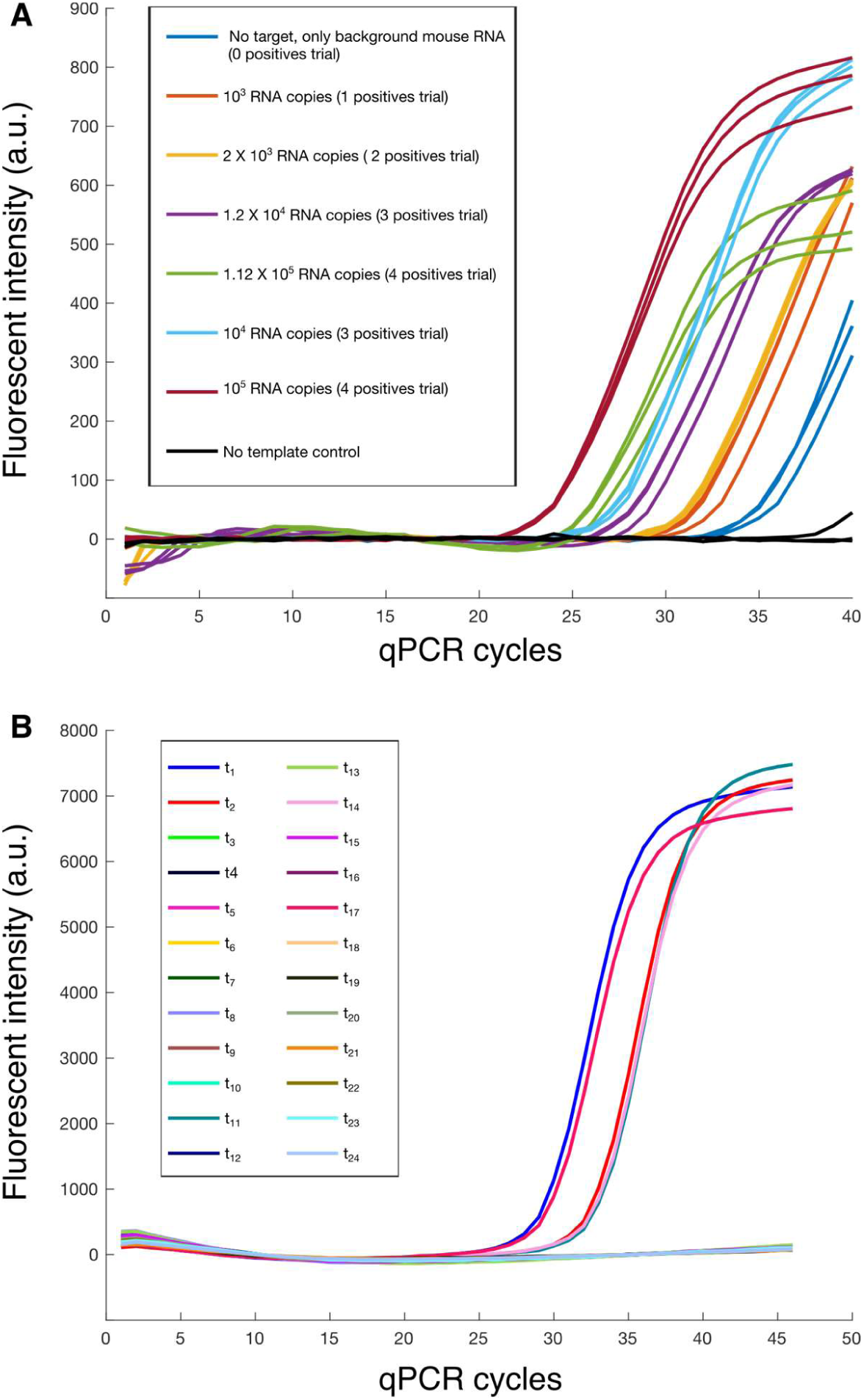
**A**. Amplification curves (Fluorescent intensity vs cycle number) for the tests performed to validate the 16 × 40 matrix. The C_t_ values derived from these curves were used to emulate the values (values within the standard deviation of the mean Ct replicate value for each condition) in the 16 × 40 matrix. The C_t_ values emulated are depicted in Table 2. **B**. Amplification curves corresponding to pooled tests for the 24 × 60 matrix. The corresponding C_t_ values are in Table 4.

**Table 3.**
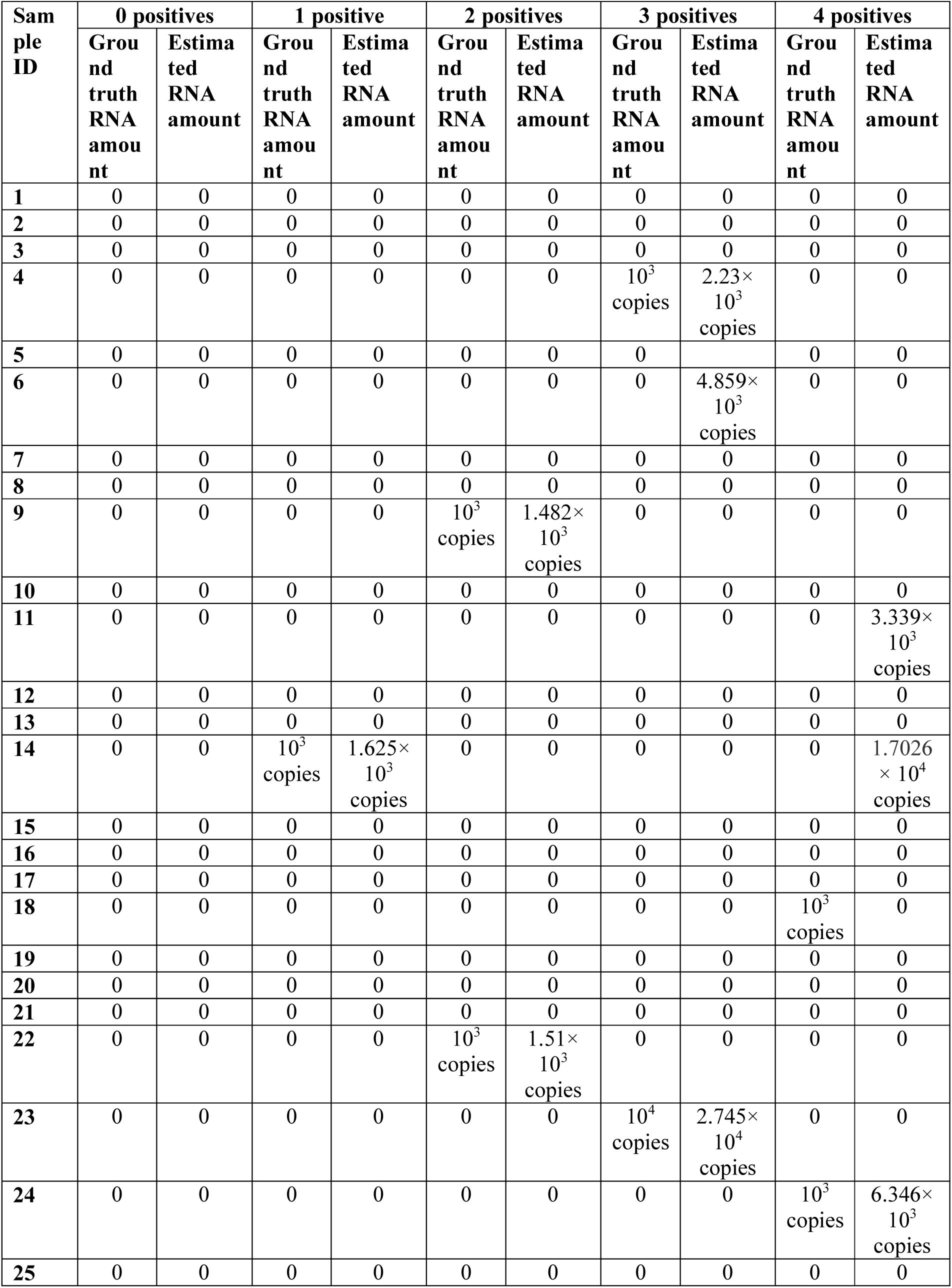
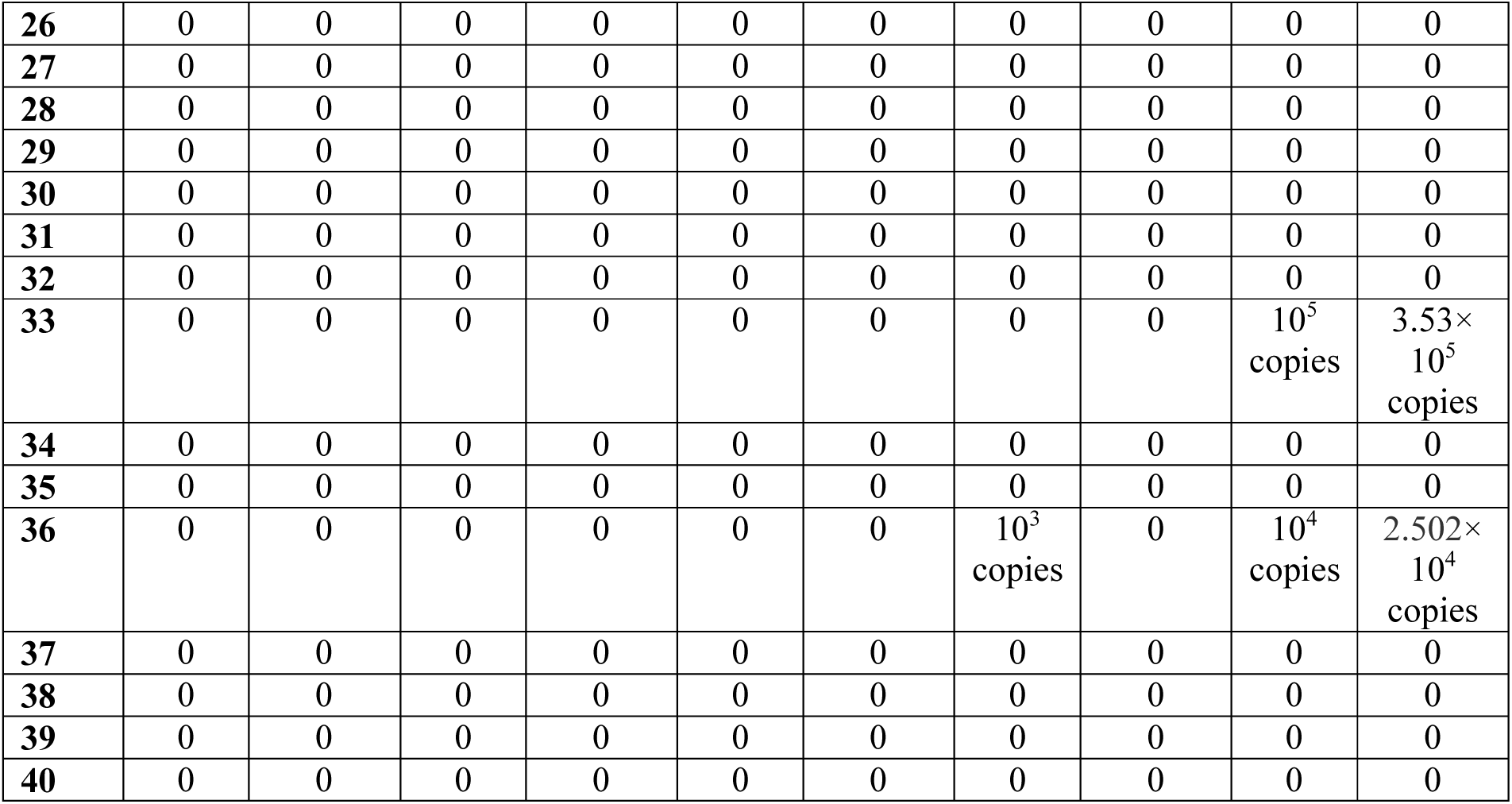
Ground truth RNA amounts for each of the 40 samples compared to the RNA amounts estimated using the Tapestry decoding algorithm.

24 × 60 pooling matrix: Table 3 shows the cycle threshold (C_t_) values obtained for the 24 RT-qPCR tests corresponding to a trial with 2 positive samples out of a total of 60 samples. Fig 2B shows the corresponding amplification curves. Table 4 shows the ground truth DNA amounts for each of the 60 samples and compares them to the values estimated using the Tapestry decoding algorithm. The algorithm correctly identified the status of all samples with zero false negatives and zero false positives.

**Table 4.** Cycle thresholds (C_t_) values for the 24 × 60 pooling matrix from Fig 1C. See experimental methods section for details on experimental setup. Amplification curves are shown in Fig. 2B.

| Test ID | 2 positive samples |
| --- | --- |
| t1 | 29.81 |
| t2 | 32.97 |
| t3 | Threshold not reached |
| t4 | Threshold not reached |
| t5 | Threshold not reached |
| t6 | Threshold not reached |
| t7 | Threshold not reached |
| t8 | Threshold not reached |
| t9 | Threshold not reached |
| t10 | Threshold not reached |
| t11 | 33.41 |
| t12 | Threshold not reached |
| t13 | Threshold not reached |
| t14 | 33.282 |
| t15 | Threshold not reached |
| t16 | Threshold not reached |
| t17 | 30.223 |
| t18 | Threshold not reached |
| t19 | Threshold not reached |
| t20 | Threshold not reached |
| t21 | Threshold not reached |
| t22 | Threshold not reached |
| t23 | Threshold not reached |
| t24 | Threshold not reached |

**Table 5.** Ground truth DNA amounts for each of the 60 samples compared to the DNA amounts estimated using the Tapestry decoding algorithm.

| Sample ID | Ground truth DNA amount | Estimated DNA amount | Sample ID | Ground truth DNA amount | Estimated DNA amount |
| --- | --- | --- | --- | --- | --- |
| P1 | 0 | 0 | P31 | 0 | 0 |
| P2 | 0 | 0 | P32 | 0 | 0 |
| P3 | 0 | 0 | P33 | 0 | 0 |
| P4 | 0 | 0 | P34 | 0 | 0 |
| P5 | 0 | 0 | P35 | 0 | 0 |
| P6 | 0 | 0 | P36 | 0 | 0 |
| P7 | 0 | 0 | P37 | 0 | 0 |
| P8 | 0 | 0 | P38 | 0 | 0 |
| P9 | 0 | 0 | P39 | 0 | 0 |
| P10 | $10^3$ copies | $1.14 \times 10^3$ copies | P40 | 0 | 0 |
| P11 | 0 | 0 | P41 | 0 | 0 |
| P12 | 0 | 0 | P42 | 0 | 0 |
| P13 | 0 | 0 | P43 | 0 | 0 |
| P14 | 0 | 0 | P44 | 0 | 0 |
| P15 | 0 | 0 | P45 | 0 | 0 |
| P16 | 0 | 0 | P46 | 0 | 0 |
| P17 | 0 | 0 | P47 | 0 | 0 |
| P18 | 0 | 0 | P48 | 0 | 0 |
| P19 | 0 | 0 | P49 | 0 | 0 |
| P20 | 0 | 0 | P50 | 0 | 0 |
| P21 | 0 | 0 | P51 | 0 | 0 |
| P22 | 0 | 0 | P52 | 0 | 0 |
| P23 | 0 | 0 | P53 | 0 | 0 |
| P24 | 0 | 0 | P54 | 0 | 0 |
| P25 | 0 | 0 | P55 | 0 | 0 |
| P26 | 0 | 0 | P56 | 0 | 0 |
| P27 | 0 | 0 | P57 | 0 | 0 |
| P28 | $10^4$ copies | $9.753 \times 10^3$ copies | P58 | 0 | 0 |
| P29 | 0 | 0 | P59 | 0 | 0 |
| P30 | 0 | 0 | P60 | 0 | 0 |

## Related Work

Group testing and its application to medical diagnosis by pooling goes back to a 1943 paper by Dorfman [6]. The application of compressed sensing to group testing in the presence of noise was introduced by Atia and Saligrama [10]. Two exciting preprints proposing applications of compressed sensing ideas to COVID-19 testing with real-time RT-qPCR have appeared online in the last few days [3,5]. Similar to our work, both these approaches argue that nonadaptive pooling has substantial advantages over adaptive pooling in the regime of low prevalence rates by enabling single-round testing of many samples with fewer tests.

While [3] is a theoretical proposal, [5] has put out proof-of-concept studies with COVID19 samples. We now compare our scheme with the schemes proposed in [3, 5], and point out some possible advantages our scheme may have.

1. Sparse Pooling Matrix: The pooling matrix A in [5] is based on Reed-Solomon error correcting codes. The pooling matrix in [3] is based on random matrices. Our pooling matrix is based on combinatorial designs known as Steiner triples. Each sample in our scheme goes to 3 pools, as opposed to 6 pools in [5] which cuts down our pipetting time by half. In simulation experiments, we also find our matrices performing better than the other two in terms of sensitivity and specificity.
2. Noise model: We propose an explicit noise model where cycle thresholds are measured with additive Gaussian noise, leading to a multiplicative noise on the reconstructed vector y. Such explicit noise models are not considered in [3,5].
3. Bespoke reconstruction algorithm: Because of the unique combination of sparse 0-1 pooling matrix, multiplicative noise model, nonnegative x vector, and one-sided error seen in this problem, we have modified existing reconstruction algorithms and obtained a corresponding improvement in performance.
4. Sparsity estimator and graceful failure: Any pooled testing scheme that takes prevalence rate as input runs the risk that the input prevalence rate may grossly underestimate the true prevalence rate, especially as the disease progresses in a population. It is important to design the scheme so that in such settings, it can (a) recognize that it is operating in a regime where the true prevalence rate exceeds the prevalence rate it was designed for, (b) maintain very high sensitivity of the test, (c) return a list of “suspected positives” and an estimate of how many of these suspected positives are truly positives. Our algorithm is able to achieve all three objectives. We employ a sparsity estimator that achieves (a). We design our recovery algorithm to guarantee (b) and achieve (c).
5. Deployment challenges: It is clear that nonadaptive strategies, for all their advantages, are in general more complex than simple pooling strategies. This raises real questions about whether they can be deployed effectively in the real world with so many moving parts. A big part of answering such a question is coming up with a way to encapsulate the complexity so that at the end of the testing laboratory the protocol to perform is made as simple as possible. We have achieved this with our Android app BYOM which gives clear instructions for pippetting into pools. It receives cycle threshold values, and solves for the viral loads in the cloud, and returns a list of the positive, suspected positive, and negative samples.

## Discussion

We have demonstrated a method for creating multiple pools from a given set of samples, in such a way that by testing the pools in a single round of RT-qPCR we can reconstruct the individual viral loads of each sample with high sensitivity and specificity. The method is designed to produce zero false negatives, and with a false positive rate that can be traded off with the overall prevalence rate that the method can handle. In general, the method works best for low prevalence rates (1-10% depending on the number of samples). For 40 and 60 samples with up to 2 positives, we have shown using mock samples (a 1kb single stranded RNA fragment and a circular DNA plasmid containing the complete nucleocapsid gene from the SARSCoV-2 virus) that the method works with zero false negatives and zero false positives. If the prevalence rate happens to be higher than expected, for example with 40 samples and 3 or 4 positives, the method fails gracefully - that is, it still provides sure positives, sure negatives and unsure positives. Moreover, the method can suggest the minimum number of new tests that need to be done to resolve the unsure cases - thus the information from the first tests is not wasted.

Our experimental and theoretical validation of the method suggest several use cases for Tapestry pooling:

1. **Single PCR run which tests 105 samples using 45 tests (pool size 8)** Currently a single 96-well PCR run can only test 46 individuals. Tapestry pooling thus provides more than 100% saving, with zero false negatives and vanishingly small false positives for upto 10% prevalence rate. Safe enough for treatment to be based on the test.
2. **Single PCR run which tests 195 samples using 45 tests (pool size 9)** This works for prevalence rates upto 5% with a single PCR run, providing zero false negatives and slightly higher false positives. This would therefore be appropriate for screening of asymptomatic subjects, such as Hospital Staff, CISF guards at airports, police personnel, delivery persons, entire apartment blocks where one person is found positive - other high-risk populations who are potential super-spreaders can be tested everyday.
3. **Two parallel PCR runs which test 399 samples using 63 test (pool size 13)** This is appropriate for larger groups of asymptomatic subjects, such as whole Neighborhoods or large hospitals or passengers arriving at an airport. It requires two technicians for pipetting. False negatives remain zero, and false positives are small for prevalence rates upto 2.5%. If the prevalence rate is higher the algorithm has a graceful failure mode.
4. **Two PCR runs which test 961 samples using 93 tests (pool size 32)** This provides massive savings in kits, RNA and PCR effort and is appropriate for populations with low prevalence rate of around 1%, for example passengers arriving at an airport, or testing in high population density neighborhoods. The larger amount of pipetting would require liquid handling robots. The pool size is also bigger, and experiments with real samples are needed to quantify the false negatives that might arise from such large dilution.

As mentioned in the last use case, experiments are required with real samples to quantify the extent to which dilution in large pools introduces false negatives, which is necessary for calibration of our quantitative method for real-world cases, and to validate the method for clinical samples using a double-blind protocol. These experiments are ongoing.

## Data Availability

All the data referred to in the manuscript is available in the manuscript. Installation files for the phone app are made available at the google drive link below. The installer will need to activate “Developer mode” on their Android device before installation.

https://play.google.com/store/apps/details?id=com.app.byom

## Acknowledgements

MG was supported by SERB grants EMR/2017/004089 and MTR/2018/000817. SK thanks the Simons Foundation for funding. NCBS authors thank the Department of Atomic Energy, Government of India, for funding under project no. 12-R&D-TFR-5.04-0800. SV thanks Wellcome Trust-DBT Alliance Early Career Fellowship (IA/E/16/1/502996) for funding. We thank Arati Ramesh, Kirti Gupta, Satyajit Mayor, Jaikumar Radhakrishnan for useful discussions.

## Protocol for Tapestry Pooling

**Note:** This protocol has been validated with in vitro experiments that used synthetic RNA and DNA fragments and additionally, its expected behavior has been confirmed using computer simulations. Validation with clinical samples is ongoing. We are looking for clinical collaborators with access to patient samples. Please contact the corresponding author if you wish to validate this protocol on clinical samples.

Equipment and consumables
- Vortex mixer
- Plate centrifuge
- Micropipettes (2 or 10 μL, 200 μL and 1000 μL)
- 12 channel multichannel micropipettes (1-10 μl)
- Racks for 1.5 mL microcentrifuge tubes
- 96-well cold blocks
- Molecular grade water, nuclease-free
- 10% bleach (1:10 dilution of commercial 5.25-6.0% hypochlorite bleach)
- DNAZap (Ambion, cat. #AM9890) or equivalent
- RNAse Away (Fisher Scientific; cat. #21-236-21) or equivalent
- Disposable powder-free gloves and surgical gowns
- Aerosol barrier pipette tips
- 1.5 mL microcentrifuge tubes (DNase/RNase free)
- 0.2 mL, 96 well PCR reaction plates with sealable covers (DNase/RNase free)
- An Android phone or tablet with the BYOM Smart Testing application installed on it (henceforth referred to as “the app”). This can be downloaded for free from the Google Playstore link: https://play.google.com/store/apps/details?id=com.app.byom

Reagents
- Nasopharyngeal swabs stored in Viral Transport Medium (henceforth referred to as sample.)

Warnings and Precautions (adapted from the United States Center for Disease Control)
- All patient specimens and positive controls should be considered potentially infectious and handled accordingly.
- Do not eat, drink, smoke, apply cosmetics or handle contact lenses in areas where reagents and human specimens are handled.
- Handle all specimens as if infectious using safe laboratory procedures.
- Specimen processing should be performed in accordance with national biological safety regulations.
- Use personal protective equipment such as (but not limited to) gloves, eye protection, and lab coats while performing this assay and handling materials including samples, reagents, pipettes, and other equipment and reagents.
- Amplification technologies such as PCR are sensitive to accidental introduction of PCR product from previous amplification reactions. Incorrect results could occur if either the clinical specimen or the real-time reagents used in the amplification step become contaminated by accidental introduction of amplification product (amplicon). Workflow in the laboratory should proceed in a unidirectional manner.

- Maintain separate areas for assay setup and handling of nucleic acids.
- Change aerosol barrier pipette tips between all manual liquid transfers.
- During preparation of samples, compliance with good laboratory techniques is essential to minimize the risk of cross-contamination between samples, and the inadvertent introduction of nucleases into samples during and after the extraction procedure. Proper aseptic technique should always be used when working with nucleic acids.
- Maintain separate, dedicated equipment (e.g., pipettes, microcentrifuges) and supplies (e.g., microcentrifuge tubes, pipette tips) for assay setup and handling of extracted nucleic acids.
- Wear a clean lab coat and powder-free disposable gloves (not previously worn) when setting up assays.
- Change gloves between samples and whenever contamination is suspected.
- Keep reagent and reaction tubes capped or covered as much as possible.
- Work surfaces, pipettes, and centrifuges should be cleaned and decontaminated with cleaning products such as 10% bleach, “DNAZap™” or “RNase AWAY®” to minimize risk of nucleic acid contamination. Residual bleach should be removed using 70% ethanol.

### Protocol

<u>Note: It is important to avoid contamination by strictly following the steps listed in Warnings and Precautions above.</u>

1. Perform RNA isolation from the nasopharyngeal swabs using approved RNA extraction kits and protocols. The isolated RNA may be concentrated using a speed-vac to increase the sensitivity of the test. (Alternatively, this step could be done after pooling samples that have been deactivated in lysate buffer. In this case, perform this step after step 6 and before step 7.)
2. Open the BYOM Smart Testing app, and select **Start New Test** from the home screen.

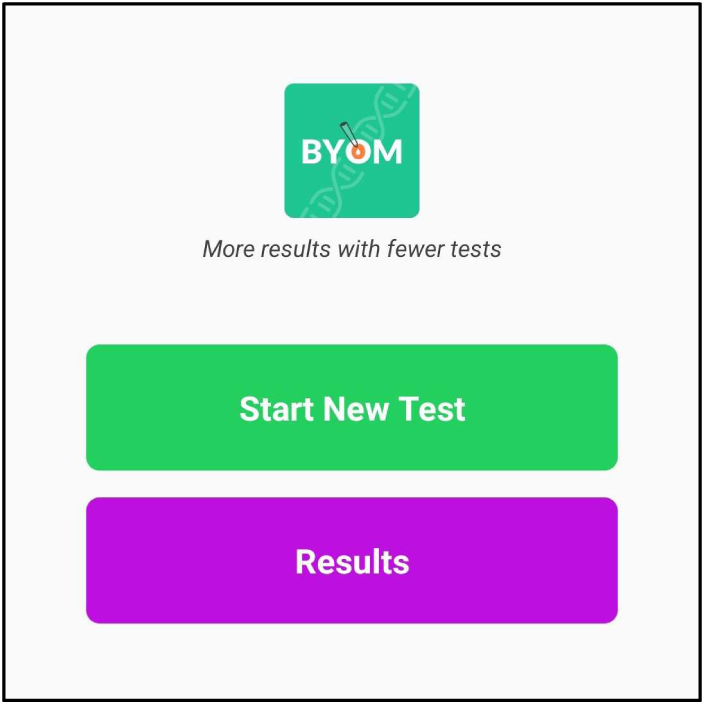
3. Enter a unique name for the test (include date and time in the name for unique identification).

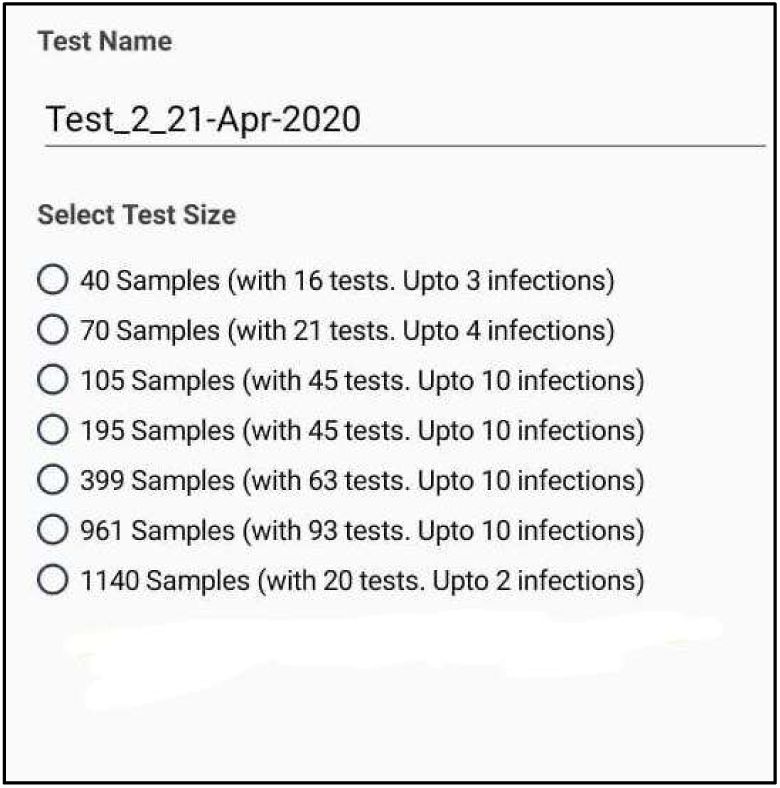
4. Select a test size based on number of samples and number of positives expected. Click next to go to the screen with pipetting instructions. The number of samples indicates the number of samples to be tested and the number of tests indicates the number of pooled Real-Time RT-PCR tests that will be performed.
5. Place an empty 0.2mL 96 well PCR reaction plate (or individual 0.2ml tubes) on a 96-well cold block/ ice and prepare to pool samples into its wells. Serially number the samples as Sample 001, Sample 002, etc.
6. Starting from Sample 001, the app will instruct the user to pipette the RNA extracted from the patient samples into specified wells (indicated by flashing orange dots) of the PCR reaction plate. The grey wells indicate all the wells that will be used for the pools You may pipette any volume (we recommend between 2uL and 10uL) as long as you are consistent across all samples and have enough pooled volume in each well to perform the downstream Real Time RT-PCR test. Clicking the **NEXT** button at the bottom of the screen or swiping to the left brings up instructions for the next sample to be pipetted, while clicking **Previous** or swiping to the right brings up instructions for the previous sample that was pipetted. Alternatively, pipetting instructions are also available on the app as downloadable PDFs.

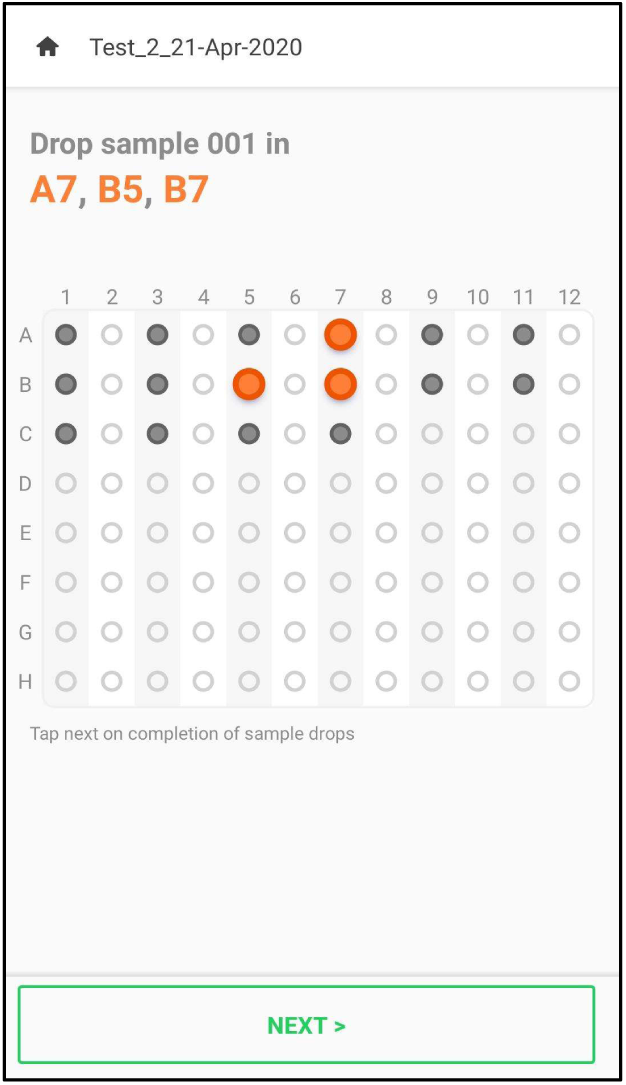
7. Once all samples have been pipetted, the app will return to the home screen and show a success message (E.g. **40 Samples Collected**). The PCR reaction plate must then be tightly sealed to prevent sample loss and spun down for 30 seconds in a plate centrifuge to collect all liquid at the bottom. The PCR reaction plate must then be gently vortexed for 5 seconds to ensure the liquid in each well is uniformly mixed. Finally, the PCR reaction plate is again spun down for 30 seconds in a plate centrifuge to collect all liquid at the bottom.
8. The pooled RNA samples in the PCR reaction plate are now ready for Real-Time RT-PCR tests. Perform all steps (RNA extraction, PCR setup etc.) according to the instructions of the Real-Time RT-PCR diagnostic panel that you will use, but using pooled samples instead of individual samples. In a typical PCR reaction, 5ul of the pooled samples will be used in a 25ul RT-PCR reaction.
9. Once the results of the Real-Time RT-PCR tests are available, collect all the C_t_ values and prepare to enter them into the app. If you performed a multiplexed PCR with multiple genes tested, enter Ct values for each gene that is testing for COVID-19. Don’t enter Ct values for the positive control RP gene into the app, but verify it at your end as per RT-PCR protocol steps.
10. Open the app and on the homescreen click **Results**. Find the corresponding test from the list of recorded tests and click **Input C_t_ values**. For each pooled test that was positive, uncheck the check box that says “Threshold not reached.” This brings up a numerical entry box where you can enter the Ct value (For multiplexed tests, this will bring up such a screen for every gene tested. This feature is not online in the app at the time of writing this protocol.)

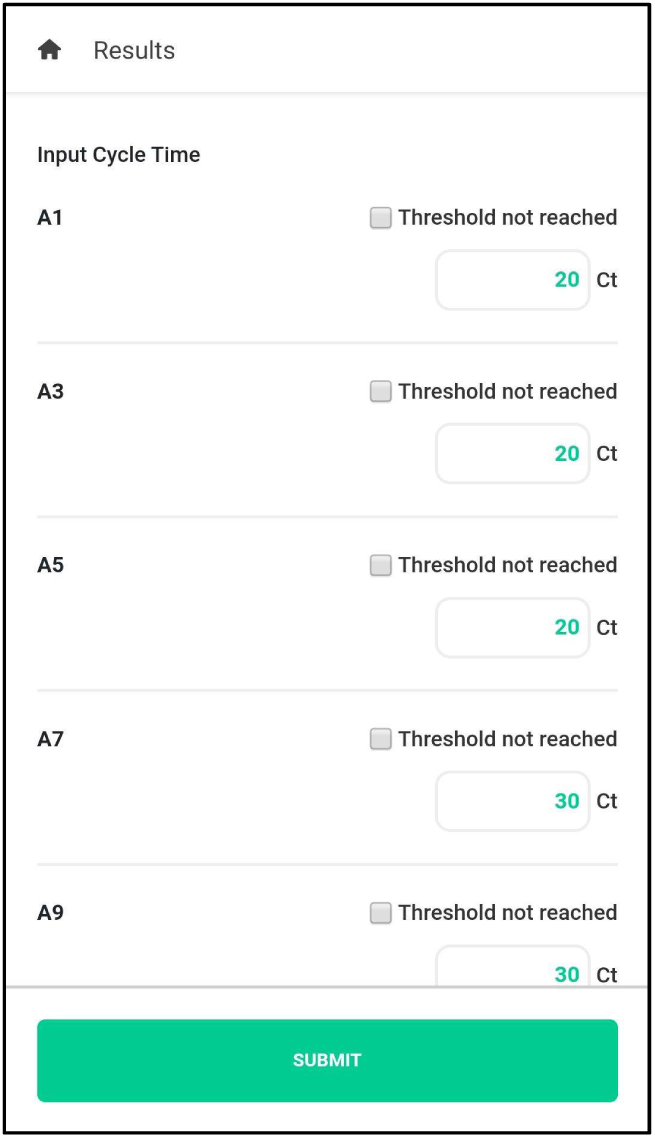

For a test that was negative, you don’t need to uncheck the check box. Click submit. The app will decode and display results for each individual sample. A list of positive samples, and a list of “possibly positive samples” will be displayed. All other samples are negative.

